# Trends and associated factors for Covid-19 hospitalisation and fatality risk in 2.3 million adults in England

**DOI:** 10.1101/2021.11.24.21266818

**Authors:** T Beaney, AL Neves, A Alboksmaty, K Flott, A Fowler, JR Benger, P Aylin, S Elkin, A Darzi, J Clarke

**Author notes:** Corresponding Author: Dr Thomas Beaney, Patient Safety Translational Research Centre, Institute of Global Health Innovation, Imperial College London, London, SW7 2AZ, United Kingdom.

## Abstract

**Background:** The Covid-19 case fatality ratio varies between countries and over time but it is unclear whether variation is explained by the underlying risk in those infected. This study aims to describe the trends and risk factors for admission and mortality rates over time in England.

**Methods:** In this retrospective cohort study, we included all adults (≥18 years) in England with a positive Covid-19 test result between 1^st^ October 2020 and 30^th^ April 2021. Data were linked to primary and secondary care electronic health records and death registrations. Our outcomes were i) one or more emergency hospital admissions and ii) death from any cause, within 28 days of a positive test. Multivariable multilevel logistic regression was used to model each outcome with patient risk factors and time.

**Results:** 2,311,282 people were included in the study, of whom 164,046 (7.1%) were admitted and 53,156 (2.3%) died within 28 days. There was significant variation in the case hospitalisation and mortality risk over time, peaking in December 2020-February 2021, which remained after adjustment for individual risk factors. Older age groups, males, those resident in more deprived areas, and those with obesity had higher odds of admission and mortality. Of risk factors examined, severe mental illness and learning disability had the highest odds of admission and mortality.

**Conclusions:** In one of the largest studies of nationally representative Covid-19 risk factors, case hospitalisation and mortality risk varied significantly over time in England during the second pandemic wave, independent of the underlying risk in those infected.

## Background

The Covid-19 case fatality ratio (CFR) varies widely between countries^1^ and definitions of mortality differ across the world, making comparisons challenging.^2^ In England, the most widely reported measure is mortality within 28 days of a positive test.^3^ Up to 21 September 2021, 539,921 hospital admissions and 118,846 deaths have occurred in England, out of a total of 6,398,633 cases, giving a crude case hospitalisation ratio (CHR) of 8.4% and a CFR of 1.9%.^4^ Previous epidemiological studies have shown variation in the CFR over time,^1,5^ but without individual level data it is unclear the extent to which this variation is accounted for by differences in the risk of those infected.

Many risk factors for death from Covid-19 have been characterised, such as increased age, male gender, and obesity.^6^ Several long-term conditions are strongly linked to a higher mortality risk; in England, this led to the early adoption of a ‘Clinically Extremely Vulnerable’ (CEV) status for those deemed to be at highest risk, subsequently advised to isolate to reduce transmission.^7^ Previous studies have focussed on the ‘first wave’ of the pandemic in the first half of 2020, which may not be representative of subsequent pandemic waves, particularly given advances in the management of Covid-19 patients and the emergence of new variants.^8^ Furthermore, to our knowledge, no study to date has used data with national coverage, including all laboratory confirmed Covid-19 test results linked to electronic health record (EHR) data.

The main aim of this paper is to describe the changing trends in the Covid-19 case hospitalisation risk (CHR) and case fatality risk (CFR) in England over time, during the ‘second wave’ of the pandemic (i.e., from 1^st^ October 2020 to 30^th^ April 2021). The secondary aims are to identify patient characteristics associated with hospitalisation and mortality risk; and to evaluate whether residual unexplained variation in the CHR and CFR remains after accounting for differences in the underlying risk factors of those infected.

## Methods

### Study design and population

We conducted a retrospective cohort study including all adults (≥18 years) resident in England with a positive Covid-19 test result from 1st October 2020 to 30th April 2021, excluding people resident in care homes. Study participants were followed-up for 28 days from the date of a first positive test. The two primary outcomes were i) one or more emergency hospital admissions and ii) death from any cause within the 28 days of the positive test.

### Data sources and data processing

Several datasets were linked for this study and provided by NHS Digital as part of an evaluation of the NHS England Covid Oximetry @home programme.^9^ Covid-19 testing data was sourced from the Public Health England (PHE) Second Generation Surveillance System,^10^ which is the national laboratory reporting system for positive Covid-19 tests, covering the period from 1^st^ October 2020 to 30th April 2021. Primary care data came from the General Practice Extraction Service (GPES) Data for Pandemic Planning and Research (GDPPR).^11^ Data on hospital admissions came from Hospital Episode Statistics (HES) data set up to 31^st^ May 2021, linked to Office for National Statistics (ONS) data on death registrations up to 5^th^ July 2021. Datasets were linked using a de-identified NHS patient ID. Participants who could not be linked from testing data to at least one of GDPPR or HES were excluded.

Patient demographics were derived from GDPPR, or where missing, from HES. Lower layer super output area (LSOA) of residence was linked to indices of relative deprivation using deciles of Index of Multiple Deprivation (IMD) 2019.^12^ Residence in a care home, CEV status, body mass index (BMI) and smoking status were derived from GDPPR only. Chronic conditions were extracted from GDPPR based on Systematised Nomenclature of Medicine Clinical Terms (SNOMED-CT) codes pertaining to relevant diagnosis code clusters. Only codes recorded prior to the date of a positive Covid-19 test were included, to exclude any diagnoses that might have resulted from Covid-19 infection. Where the latest code indicated resolution of a condition, the diagnosis was excluded for that individual. Further details on data curation, as well as a list of all included SNOMED-CT codes, are provided in Supplementary Appendix A.

### Statistical analysis

Patients were followed from date of first positive Covid-19 test to emergency hospital admission or death within 28 days. Mixed effects logistic regression was conducted for each outcome, with a two-level hierarchical model incorporating Clinical Commissioning Group (CCG) of residence as a random intercept. Time, represented by week of Covid-19 test, was modelled as a restricted cubic spline with five knots placed at equally spaced percentiles.^13^ Two models were run for each outcome:

1. Model 1: incorporating age category and time splines along with their interaction
2. Model 2: incorporating age category and time splines along with their interaction and including all additional patient level covariates: sex, ethnicity, IMD decile, BMI category, CEV status, smoking status, and presence of chronic conditions.

For model 2, multiple imputation using chained equations was used to impute missing values of covariates, under the assumption that values were missing at random. All variables included in the analysis model were included in the imputation model.^14^ Fifteen imputations were created, with a burn-in of 10 iterations which gave adequate precision and convergence, respectively (see Supplementary Appendix B). A sensitivity analysis was performed using complete cases only. Hosmer-Lemeshow plots of predicted against observed probabilities for each decile of predicted probability indicated adequate calibration (Supplementary Appendix B, Figures S1 and S2).

For each outcome, the predicted probability of the outcome was computed within each age group and study week stratum to calculate age- and time-specific case hospitalisation risk (CHR) and case fatality risk (CFR). These were calculated using the fixed portion of the model (assuming zero random effects). The relative changes in the CHR and CFR over time were calculated as the predicted probability in each week relative to the first study week (1st-4th October 2020) in each age group. In adjusted models (model 2), other model covariates were set to the population mean (or proportion for categorical variables) within each age group. For CEV status, an additional sub-analysis was conducted adjusting only for age category and time splines (and their interaction), sex, ethnicity and IMD decile. Further details of the statistical methods are given in Supplementary Appendix B.

Analyses were conducted in the Big Data and Analytics Unit Secure Environment, Imperial College, using Python version 3.9.5 and Stata version 17.0 (StataCorp). The work was conducted as part of a wider service evaluation, approved by Imperial College Healthcare Trust on December 3^rd^ 2020. Data access was approved by the Independent Group Advising on the Release of Data (IGARD; DARS-NIC-421524-R0Y3P) on April 15^th^ 2021.

## Results

From 1st October 2020 to 30th April 2021, data were available for 2,433,768 individuals with a positive Covid-19 test result in England. Data for 34,317 (1.4%) participants with a positive test result could not be linked to either primary or secondary care records and were excluded. Care home residents accounted for 3.7% of the total (n=88,169) and were excluded from further analyses, resulting in a total population of 2,311,282.

Characteristics of the study population are provided in Table 1. The mean (SD) age of participants was 44.3 (17.1) years, with 43.6% under 40 years. The majority were female (54.3%) and of white ethnicity (72.8%). There were relatively higher proportions from more deprived deciles of IMD, with 56.7% in the bottom five deciles. Similar proportions of subjects with a healthy weight (28.4%), overweight (28.1%) or obese (26.1%) were observed, and only 3.4% were underweight. 16.3% were current smokers and 8.3% were designated as CEV. Chronic respiratory disease (21.2%), hypertension (15.0%) and diabetes (8.6%) were the three most prevalent chronic conditions in the population.

**Table 1:** Characteristics of the study population (N=2,311,282) with hospital admissions and deaths within 28 days.

|  | Total |  | Hospital admissions |  | Deaths |  |
| --- | --- | --- | --- | --- | --- | --- |
|  | Number | Percentage | Number | Percentage | Number | Percentage |
| **Age category (years) and CEV status** |  |  |  |  |  |  |
| Mean (SD) | 44.3 (17.1) years |  |  |  |  |  |
| 18-39 | 1,007,474 | 43.6% | 19,834 | 2.0% | 429 | 0.1% |
| 40-49 | 442,337 | 19.1% | 18,897 | 4.3% | 989 | 0.7% |
| 50-59 | 434,690 | 18.8% | 30,138 | 6.9% | 3,054 | 0.7% |
| 60-69 | 229,209 | 9.9% | 30,070 | 13.1% | 7,009 | 3.1% |
| 70-79 | 112,379 | 4.9% | 31,436 | 28.0% | 14,068 | 12.5% |
| 80 or older | 85,193 | 3.7% | 33,671 | 39.5% | 27,607 | 32.4% |
| **Sex** |  |  |  |  |  |  |
| Female | 1,255,364 | 54.3% | 72,126 | 5.7% | 21,253 | 1.7% |
| Male | 1,010,045 | 43.7% | 86,135 | 8.5% | 29,574 | 2.9% |
| Missing | 45,873 | 2.0% | 5,785 | 12.6% | 2,329 | 5.1% |
| **Ethnicity** |  |  |  |  |  |  |
| White | 1,681,477 | 72.8% | 119,999 | 7.1% | 42,753 | 2.5% |
| Asian/Asian British | 304,685 | 13.2% | 21,900 | 7.2% | 4,539 | 1.5% |
| Black/African/Caribbean/Black British | 87,974 | 3.8% | 7,880 | 9.0% | 1,505 | 1.7% |
| Mixed/Multiple ethnic groups | 38,397 | 1.7% | 2,236 | 5.8% | 392 | 1.0% |
| Other ethnic group | 58,789 | 2.5% | 4,388 | 7.5% | 616 | 1.0% |
| Missing | 139,960 | 6.1% | 7,643 | 5.5% | 3,351 | 2.4% |
| **Index of Multiple Deprivation (IMD) decile** |  |  |  |  |  |  |
| 1 (most deprived) | 277,814 | 12.0% | 23,009 | 8.3% | 6,734 | 2.4% |
| 2 | 282,141 | 12.2% | 22,238 | 7.9% | 6,510 | 2.3% |
| 3 | 271,120 | 11.7% | 20,001 | 7.4% | 5,985 | 2.2% |
| 4 | 248,041 | 10.7% | 17,838 | 7.2% | 5,689 | 2.3% |
| 5 | 231,591 | 10.0% | 16,376 | 7.1% | 5,220 | 2.3% |
| 6 | 218,370 | 9.4% | 14,774 | 6.8% | 5,104 | 2.3% |
| 7 | 208,612 | 9.0% | 13,647 | 6.5% | 4,831 | 2.3% |
| 8 | 206,003 | 8.9% | 13,316 | 6.5% | 4,719 | 2.3% |
| 9 | 194,561 | 8.4% | 12,273 | 6.3% | 4,440 | 2.3% |
| 10 (least deprived) | 172,508 | 7.5% | 10,542 | 6.1% | 3,917 | 2.3% |
| Missing | 521 | <0.1% | 32 | 6.1% | 7 | 1.3% |
| **Body Mass Index** |  |  |  |  |  |  |
| Underweight | 78,684 | 3.4% | 3,149 | 4.0% | 2,063 | 2.6% |
| Healthy weight | 655,582 | 28.4% | 30,448 | 4.6% | 13,967 | 2.1% |
| Overweight | 649,641 | 28.1% | 48,161 | 7.4% | 15,652 | 2.4% |
| Obese | 603,303 | 26.1% | 67,638 | 11.2% | 17,199 | 2.9% |
| Missing | 324,072 | 14.0% | 14,650 | 4.5% | 4,275 | 1.3% |
| **Smoking status** |  |  |  |  |  |  |
| Never smoker | 1,363,771 | 59.0% | 82,668 | 6.1% | 21,963 | 1.6% |
| Ex-smoker | 475,558 | 20.6% | 53,186 | 11.2% | 21,757 | 4.6% |
| Current smoker | 376,057 | 16.3% | 21,260 | 5.7% | 7,005 | 1.9% |
| Missing | 95,896 | 4.1% | 6,932 | 7.2% | 2,431 | 2.5% |
| **Clinically Extremely Vulnerable** | 192,531 | 8.3% | 48,679 | 25.3% | 19,294 | 10.0% |
| **Co-morbidities** |  |  |  |  |  |  |
| Hypertension | 346,145 | 15.0% | 69,202 | 20.0% | 31,710 | 9.2% |
| Chronic cardiac disease | 126,133 | 5.5% | 37,775 | 29.9% | 22,878 | 18.1% |
| Chronic kidney disease | 14,492 | 0.6% | 5,485 | 37.8% | 3,553 | 24.5% |
| Chronic respiratory disease | 489,341 | 21.2% | 53,099 | 10.9% | 19,960 | 4.1% |
| Dementia | 13,552 | 0.6% | 5,111 | 37.7% | 4,800 | 35.4% |
| Diabetes | 199,495 | 8.6% | 44,280 | 22.2% | 18,293 | 9.2% |
| Chronic neurological disease<br>(including epilepsy) | 64,274 | 2.8% | 10,765 | 16.7% | 4829 | 7.5% |
| Learning disability | 11,627 | 0.5% | 1,775 | 15.3% | 485 | 4.2% |
| Malignancy or immunosuppression | 178,648 | 7.7% | 30,014 | 16.8% | 15,691 | 8.8% |
| Severe mental illness | 39,513 | 1.7% | 7,038 | 17.8% | 3,684 | 9.3% |
| Peripheral vascular disease | 14,453 | 0.6% | 5,266 | 36.4% | 3,664 | 25.4% |
| Stroke or TIA | 44,239 | 1.9% | 13,880 | 31.4% | 8,712 | 19.7% |
| **Total** | **2,311,282** |  | **164,046** |  | **53,156** |  |

### Case hospitalisation and fatality risk over time

Of the study population, 164,046 people were admitted to hospital at least once within 28 days of a positive test, giving a crude CHR of 7.1% over the seven-month period. 53,156 deaths occurred within 28 days of a positive test, giving a crude CFR of 2.3%. There were significant differences across age groups and over time for both the crude CHR and CFR, with risk peaking in December-February (Tables S1 and S2, respectively, and Figure S3 in Supplementary Appendix B).

### Factors associated with 28-day mortality and hospitalisation risk

Multiple imputation was used to impute missing data for 381,283 people. Multivariable logistic regression models were constructed for each outcome adjusting for all patient level covariates (model 2). Results for hospital admissions and mortality are presented in Figure 1 and Figure 2 (also Supplementary Appendix B, Tables S3 and S4). Males had 41% higher adjusted odds of admission (95% CI: 1.39-1.42) and 62% higher adjusted odds of mortality (95% CI: 1.58-1.65) compared to females. People of all four non-white ethnicities had higher odds of admission, and those of Asian and Black ethnicities also had higher odds of mortality compared to those of white ethnicity. People living in less deprived areas had lower odds of both admission and mortality compared to those in the most deprived areas. Compared to people of a healthy weight, those underweight had 10% higher odds of admission (95% CI: 1.05-1.14) and 99% higher odds of death (95% CI: 1.87-2.11). People who were overweight had a 24% increase in odds of admission (95% CI: 1.22-1.26) but 20% lower odds of death (95% CI: 0.77-0.82); those who were obese had 93% higher odds of admission (1.90-1.97) and 4% increased odds of death (95% CI: 1.01-1.07). Current smokers had lower odds of admission compared to non-smokers but an increase in the odds of death after adjustment.

**Figure 1:**
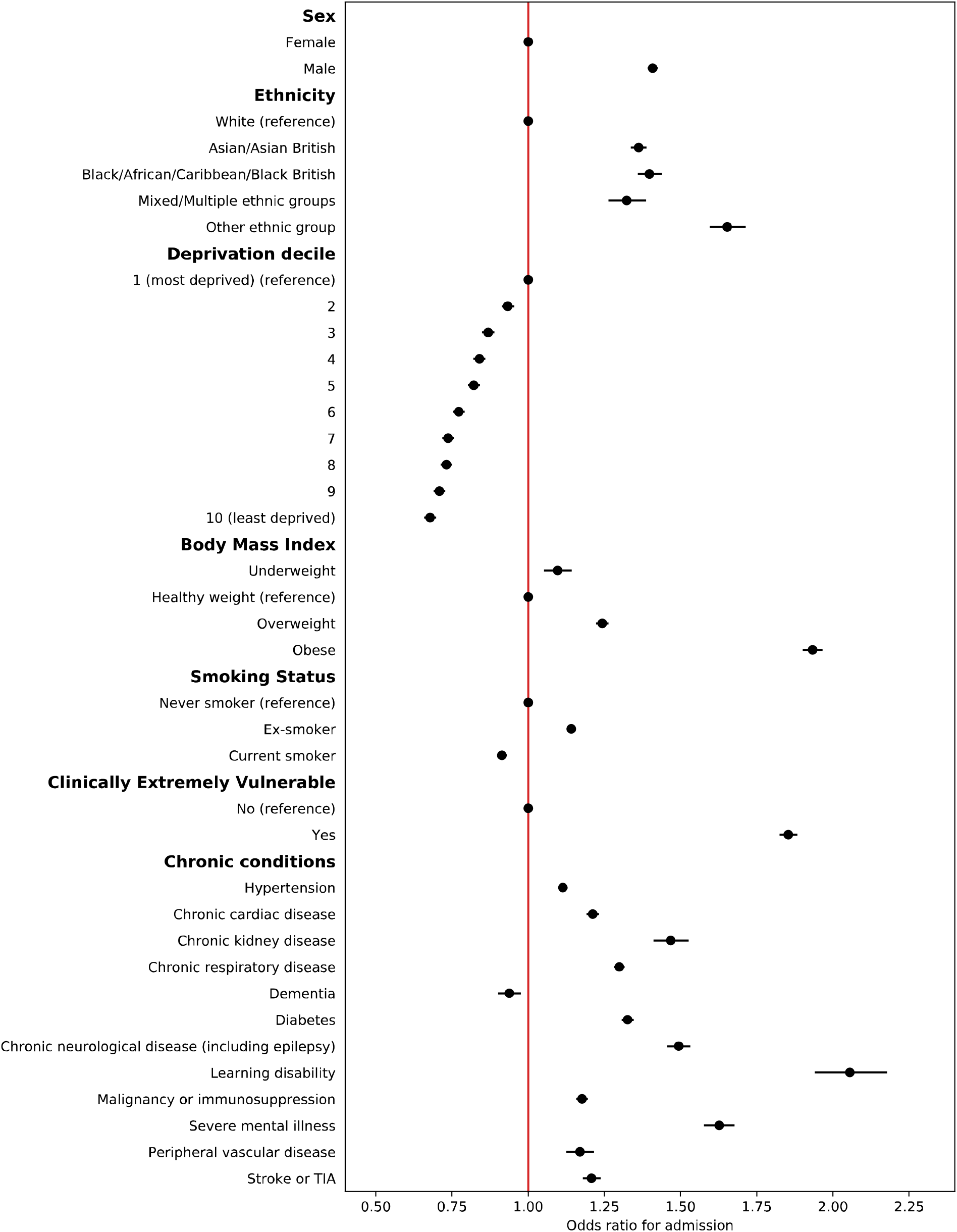
Estimated adjusted odds ratios for emergency hospital admission within 28 days for patient-level predictors from multivariable mixed effects logistic regression models (N=2,311,282) Note: circles represent estimates and bars represent 95% confidence intervals

**Figure 2:**
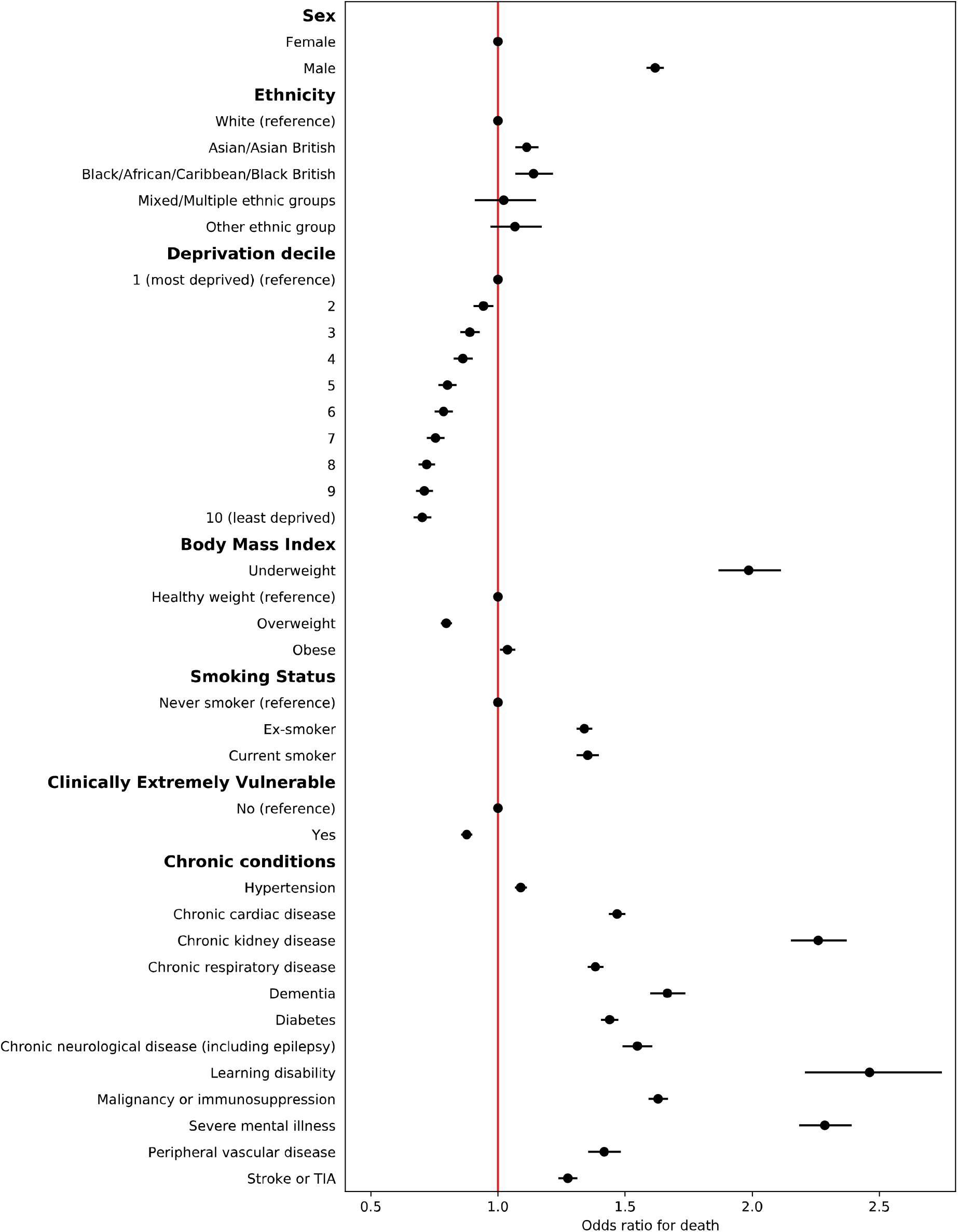
Estimated adjusted odds ratios for death within 28 days for patient-level predictors from multivariable mixed effects logistic regression models (N=2,311,282) Note: circles represent estimates and bars represent 95% confidence intervals

All chronic conditions included were strongly associated with an increase in odds of admission and death, except for dementia, which was associated with 6% lower odds of admission. People identified as CEV had 85% higher odds of being admitted to hospital (95% CI: 1.83-1.88) but 12% lower odds of death (95% CI: 0.86-0.90) after full adjustment. In a sub-analysis adjusting CEV status for age, time (and their interaction), sex, ethnicity and deprivation only, odds of admission were significantly higher (aOR 2.62, 95% CI: 2.58-2.65) as were odds of death (aOR 1.52, 95% CI: 1.49-1.55).

A sensitivity analysis of the 1,929,999 complete cases showed similar estimates to the fully adjusted model (Tables S5 and S6 in Supplementary Appendix B).

### CHR and CFR over time

A significant association remained with time for both CHR and CFR models after adjusting for all patient covariates (*p*<0.0001 in each model from likelihood ratio tests). The predicted CHR and CFR from the fully adjusted models are plotted over time and by age category in Figure 3, showing that a significant time-varying relationship remains after adjustment. The relative change in predicted CHR and CFR from the baseline predicted risk in the first week of October is shown in Figure 4 (and Figures S4 and S5 in Supplementary Appendix B). The CFR increased across all age groups, peaking between late December 2020 to early February 2021in different age groups before declining towards April. A smaller relative increase in hospitalisation risk was seen across age groups. In most age groups, CHR peaked in January, except in the 18-39 age group, which continued to increase throughout the study period.

**Figure 3:**
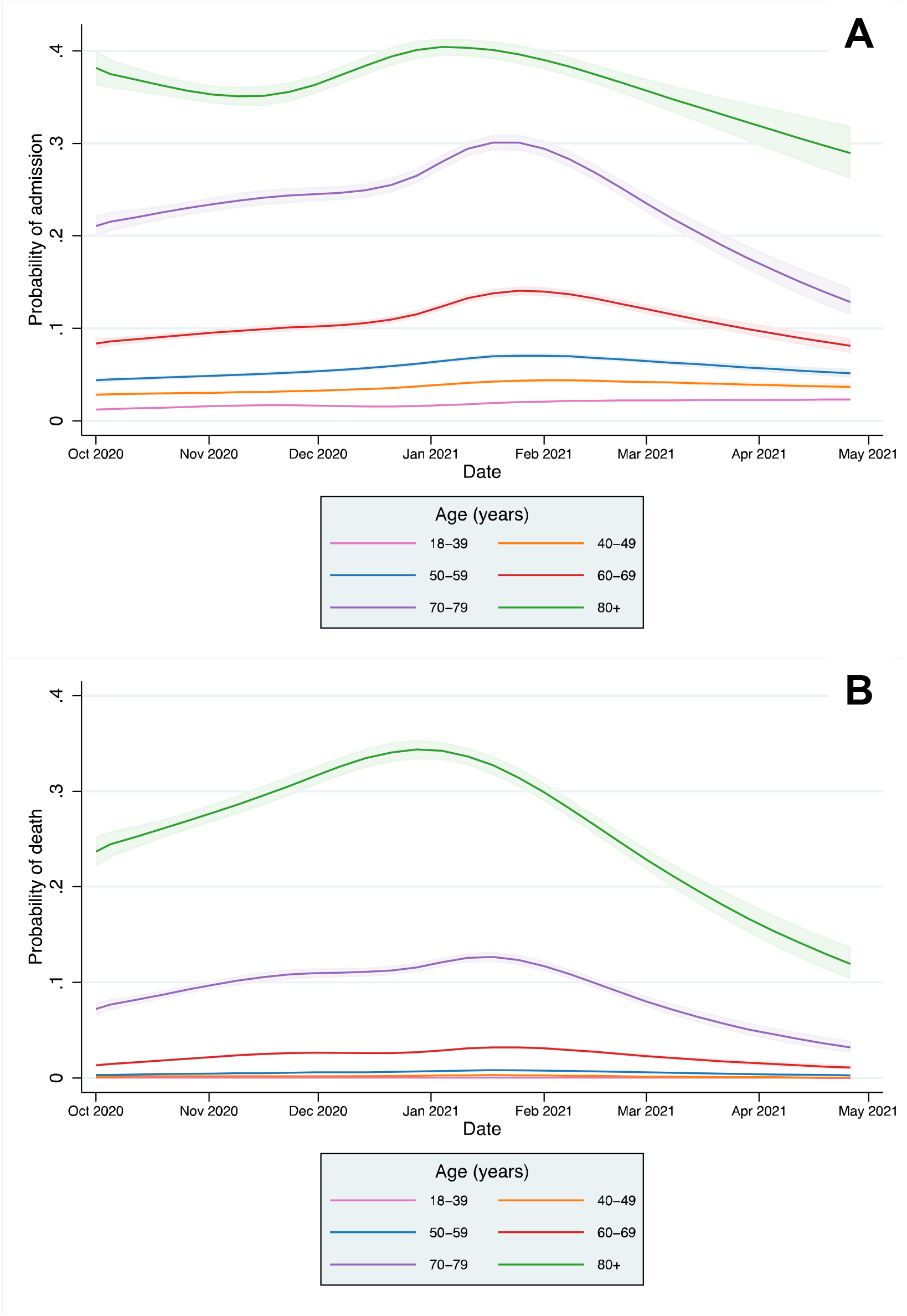
Case hospitalisation risk (CHR) (A) and fatality risk (CFR) (B) over time in people with Covid-19 from mixed effects logistic regression models, adjusted for patient-level covariates, at mean levels of each covariate (N=2,311,282) Note: shaded areas represent 95% confidence intervals

**Figure 4:**
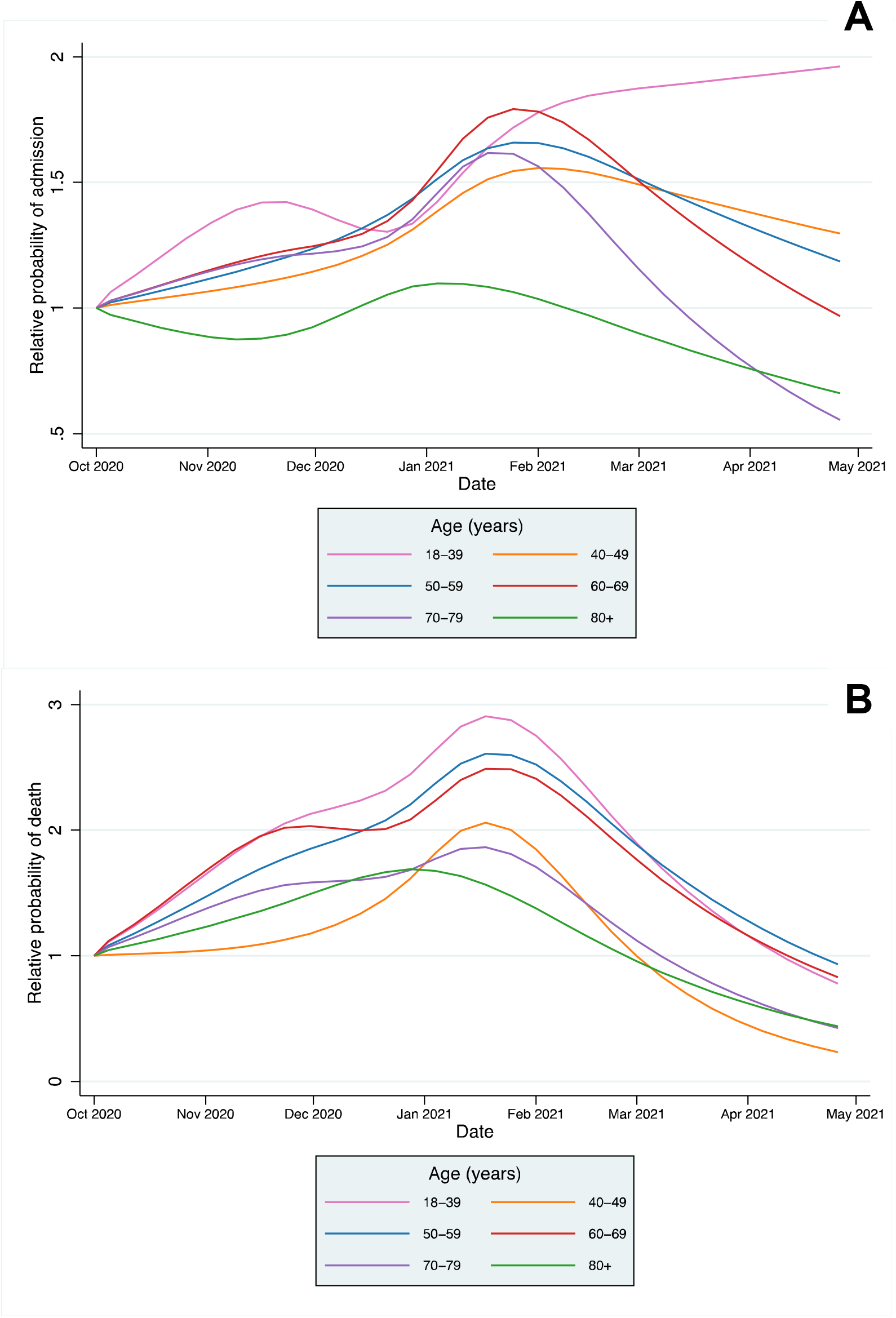
Relative change in case hospitalisation risk (CHR) (A) and fatality risk (CFR) (B) over time, compared to the first week of October, in people with Covid-19 from mixed effects logistic regression models, adjusted for patient-level covariates, at mean levels of each covariate (N=2,311,282) Note: different y axis scales for admissions and mortality

## Discussion

In this retrospective cohort study including all adults in England with a positive Covid-19 test result, there was significant variation in the 28 day CHR and CFR by age group and over time, which remained after accounting for individual risk. Demographics and chronic conditions were strongly associated with hospitalisation and death.

### Variation in CHR and CFR over time

Absolute differences in the CHR and CFR over time from 1^st^ October 2020 were greatest in older age groups, reflecting higher baseline risk, but the relative risk varied significantly across all age groups. Historically, there is a strong seasonal component to mortality in England, with figures indicating 16.8% higher mortality in winter months compared to summer months.^15^ An increased incidence of respiratory diseases, including influenza, are one of the main drivers of increased winter mortality, and the 28-day mortality metric used in this study includes deaths from non-Covid-19 causes. However, with influenza rates at lower levels than previous years, it is unlikely the variation in CFR over time can be explained by the incidence of other infectious diseases alone.^16^

Strain on the health system may also contribute to the patterns seen, with Covid-19 bed occupancy and critical care occupancy in England peaking in January 2021, associated with a lower proportion of patients seen in Accident & Emergency departments within 4 hours than in November 2020 and February 2021.^4,17^ Larger relative increases were seen in the CFR compared to the CHR, which may indicate a health system reaching full capacity and struggling to meet demand. A previous study of patients admitted to hospital with Covid-19 found a fall in mortality from March to July 2020, a time over which bed occupancy fell and evidence for new treatments, such as dexamethasone, became available.^18^ Changes to care delivery at an organisational level may also have an impact, with triage models for Covid-19 patients on the national 111 urgent care service varying between services and over time.^19^ The alpha variant (VOC-202012/01) became the dominant Covid-19 strain in England in December 2020, and has been associated with a 64% increase in 28-day mortality compared to prior variants, which may explain part of the rise in the CHR and CFR.^20^

Declines in the CHR and CFR from January 2021 onwards are likely to be explained at least partially by development of immunity, both through natural infection and by the vaccination programme, which was implemented from December 2020 in England for the highest risk cohorts.^21^ Declines in CFR and CHR are most marked in older age groups, who were the first groups eligible for vaccination. However, declines in mortality are seen across all age groups, including the 18–39 year group, the majority of whom would not have been eligible for vaccination, suggesting vaccination does not fully account for the declines observed. Availability of new treatments may also explain the falls in mortality, but not admissions, with the RECOVERY trial demonstrating the benefit of tocilizumab published in February 2021.^8,22^

### Factors associated with hospitalisation and mortality

The findings of a higher risk of mortality in males, people of Asian and Black ethnic backgrounds, and those living in more deprived areas are consistent with a previous UK cohort and confirmed in our study, including an increased risk of admission.^6^ People who were underweight were more likely to be admitted and had significantly higher risk of death, which might be partly accounted for by unmeasured associated conditions, such as frailty. People who were overweight and obese had higher risk of admission than those of a healthy weight, but mortality risk was lower in those overweight, which may indicate higher perceived risk amongst clinicians and a lower threshold for admission.

People identified as CEV were significantly more likely to be admitted but were found to have significantly lower mortality, after adjusting for other risk factors including co-morbidities. However, in partially adjusted models not including BMI, smoking, or clinical co-morbidities, those identified as CEV had significantly higher odds of death. Taken together, these findings indicate a lower threshold for clinical assessment and/or admission and escalation in CEV patients with a protective effect on mortality. All twelve included clinical co-morbidities were associated with significant increases in the odds of mortality and admission. Severe mental illness and learning disability had the strongest associations with mortality and admission, highlighting a need for more proactive care in these groups and more research into the reasons for mortality differences.^23^ Those with dementia had significantly increased odds of mortality but were less likely to be admitted, suggesting they are more likely to receive care at home, although the cohort did not include those living in care homes.

### Strengths and limitations

A strength of this study is the inclusion of routine national laboratory data for positive Covid-19 test results in adults in England with only 1.5% unable to be linked to EHR data, and as a result has lower risk of sampling bias.^24^ To our knowledge, this is the largest such study including individual level data at a national level bias. Previous studies in England on predictors of mortality are reported on a smaller cohort of patients with 40% national coverage.^6^ Use of multiple imputation assumes that data are missing at random, and we cannot rule out non-random missing patterns, particularly for data on ethnicity and deprivation, where more marginalised groups are less likely to be registered in the primary care record. However, sensitivity analyses showed inferences were similar between the complete case analysis and imputed results, suggesting limited impact of the missing data on model estimates.

Data represented here include only those who died within 28 days of a positive test result, in line with estimates reported by PHE. Deaths mentioning Covid-19 on a death certificate are an alternative metric used widely in many countries as recommended by the World Health Organisation^25^ and have tended to give a larger estimate of deaths in England, due to those attributable to Covid-19 after 28 days.^4^

Through use of linked EHR data, we were able to incorporate detailed medical factors for the study cohort. However, we were unable to explore the relationship with external factors such as Covid-19 variants. Geographical and time-varying system factors, such as proximity to a hospital and hospital capacity are likely to impact on a person’s health-seeking behaviour. However, our modelling showed only minimal residual variation accounted for by CCG level clustering (intraclass correlation coefficient <1%), suggesting these additional factors would have minimal impact on the findings. Exploring mortality risk in patients admitted to intensive care units and whether this changed over time was outside the scope of the current study but is an area for further research.

## Conclusion

Risk of hospitalisation and death from Covid-19 varied significantly over time from October 2020 to April 2021 in all age groups, independent of the underlying risk in those infected. Time-varying risks should be considered by researchers and policymakers in assessing the risks of hospitalisation and mortality from Covid-19. People with severe mental illness and learning disability were amongst those with the highest odds of both admission and mortality, indicating the need for proactive care in these groups.

## Data Availability

The patient level data used in this study are not publicly available but are available to applicants meeting certain criteria through application of a Data Access Request Service (DARS) and approval from the Independent Group Advising on the Release of Data.

## Acknowledgements

The authors would like to thank Hutan Ashrafian, Gianluca Fontana, Saira Ghafur, Melanie Leis and Mahsa Mazidi for their input and support. Data management was provided by the Big Data and Analytical Unit (BDAU) at the Institute of Global Health Innovation (IGHI), Imperial College London. The authors acknowledge support from NHS England and NHS Improvement, the National Institute for Health Research (NIHR) Imperial Biomedical Research Council (BRC) and the NIHR Imperial Patient Safety Translational Research Centre. JC acknowledges support from the Wellcome Trust (215938/Z/19/Z). The study funders did not play a role in study design; in the collection, analysis, and interpretation of data; in the writing of the report; and in the decision to submit the article for publication. In addition, researchers were independent from funders, and all authors had full access to all of the data included in this study and can take responsibility for the integrity of the data and the accuracy of the data analysis.

