## Supplementary Appendix A for "Trends and associated factors for Covid-19 hospitalisation and fatality risk in 2.3 million adults in England"

Corresponding Author:

Dr Thomas Beaney

**Data cleaning**

This section describes the cleaning process for each dataset.

*Covid-19 testing data*

Testing data was provided through the Public Health England Second Generation Surveillance System (SGSS). This dataset captures routine laboratory data on infectious diseases for England, including Covid-19, with all diagnostic laboratories required to notify positive test results within 24 hours.^1^ Data included 3,251,225 tests performed from 1^st^ October 2020 to 30^th^ June 2021, inclusive.

Provided data included test date and result date. 99% of results were reported within 5 days of the test, and 6,544 (0.2%) were reported more than 7 days from date of test. Where the result data occurred before the test date. In 1,603 cases, the result date was recorded as prior to the test date. In these instances, where the difference between the testing date and reporting date was 7 days or less, the test date and reporting date were swapped. In the 191 instances where reporting date was more than 7 days before the testing date, the test was excluded.

For this analysis, only tests performed up to 30^th^ April 2021 were included (after swapping test and result dates where applicable), given that secondary care data was available up until the end of May 2021. Of the 2,925,356 positive Covid-19 tests, 2,349,237 (80.3%) were from Pillar 2 testing, 561,570 (19.2%) from Pillar 1, and 14,549 (0.5%) from Pillar 4.^2^ Test type was recorded for Pillar 2 tests only, and of these, 2,248,497 (95.7%) were Polymerase Chain Reaction (PCR) tests, 100,740 (4.3%) were lateral flow tests.

In these analyses, only the first case of a positive test where more than one test was recorded for any individual (2,533,504). Any patient resident in a Clinical Commissioning Group (CCG) outside of England, or in NHS commission hub CCGs, were excluded resulting in testing population of 2,433,768 individuals.

*Primary care data*

Primary care data came from the General Practice Extraction Service (GPES) Data for Pandemic Planning and Research (GDPPR).^3^ Data included month and year of birth, sex, ethnicity, Lower Layer Super Output Area (LSOA) of residence, a marker for Clinically Extremely Vulnerable (CEV) status, and a marker for residence in a care home. LSOA was used to link to 2019 deciles of Index of Multiple Deprivation (IMD).^4^ Age was calculated from date of positive Covid-19 test, assuming a birthdate on the 15^th^ day of the month.

Entries include a date to which each journal item applies, and a date on which the journal item was recorded. The former was used in priority, but where missing, was replaced with the journal item recording date. For LSOA, CEV status and care home residence, only entries occurring up to the date of positive Covid-19 test were included. For month and year of birth, sex and ethnicity, if no entry were included prior to the date of Covid-19 test, then the earliest recorded entry after the test was included.

*Secondary care data*

Data on hospital admissions came from the Hospital Episode Statistics (HES) data set up to 31st May 2021, linked to Office for National Statistics (ONS) data on death registrations up to 5th July 2021.^5^ Entries were excluded where missing admission dates, provider Trust code, or patient deidentified ID. Where age was missing from GDPPR, it was derived from month and year of birth in HES using the same approach as for GDPPR.

Where multiple admission episodes were recorded within a spell, a single spell start and end date were created. Non-emergency hospital admissions were excluded from analyses. A binary indicator was created for any admission within 28 days of positive Covid-19 test. A second indicator was created for death (of any cause) within 28 days of positive Covid-19 test.

*Co-morbidities*

SNOMED codes were included in the GDPPR dataset pertaining to specific SNOMED code cluster reference sets provided by NHS Digital.^3^ 6,485 unique codes were identified from GDPPR. Codes were reviewed manually by authors TB and JC and removed if not relevant or assigned to the minimal number of relevant code clusters.

SNOMED reference clusters were aggregated into hierarchies of similar conditions. Codes in each higher-order cluster were then reviewed to ensure groupings of relevant codes and twelve relevant chronic disease categories were selected: hypertension, chronic cardiac disease, chronic kidney disease, chronic respiratory disease, dementia, diabetes, chronic neurological disease (including epilepsy), learning disability, malignancy/immunosuppression, severe mental illness, peripheral vascular disease and stroke/transient ischaemic attack (TIA). Categories for chronic respiratory disease, diabetes, epilepsy, malignancy/immunosuppression and severe mental illness included relevant medication codes. Broad diagnostic categories of diagnoses were chosen, as certain medications were not diagnostic of more granular diagnostic categories (for example, use of a long-acting bronchodilator/inhaled corticosteroid in both COPD and asthma) A full list of codes within each diagnostic category are available in Supplementary Appendix C.

For each patient in GDPPR, all relevant diagnostic codes prior to the study index date (date of positive Covid-19 test) were considered diagnostic. In cases where the latest SNOMED code indicated resolution of a condition (eg ‘Atrial fibrillation resolved (finding)’), then the diagnosis was excluded for that patient. SNOMED codes relating to drug codes were only included up to 2 years prior to the index date.

*BMI categorisation*

SNOMED codes for BMI were either diagnostic categories (eg ‘Body mass index 30+ - obesity (finding)’ or value codes (eg. ‘Body mass index (observable entity)’). Values were extracted and BMI was categorised according to the standard World Health Organisation classification of underweight (<18.5 kg/m^2^), healthy weight (18.5-24.9 kg/m^2^), overweight (25.0-29.9 kg/m^2^) and obese (≥30.0 kg/m^2^). Value codes outside of the range 5.0-100.0 kg/m2 were excluded. SNOMED codes which spanned more than one category (eg ‘Increased body mass index (finding)’) and child BMI categories were also excluded.

*Smoking categorisation*

Smoking status was categorised into ‘never-smoker’, ‘ex-smoker’ and ‘current smoker’ according to the latest SNOMED code prior to and including the index date. For any patient where the latest SNOMED code indicated ‘never-smoker’, but a prior record indicated active smoking, then the patient was re-categorised as ‘ex-smoker.

**References**

1.	Public Health England. Laboratory reporting to Public Health England: A guide for diagnostic laboratories. 2020.

2.	Department of Health & Social Care. COVID-19 testing data: methodology note [Internet]. 2021 [cited 2021 Oct 6];Available from: https://www.gov.uk/government/publications/coronavirus-covid-19-testing-data-methodology/covid-19-testing-data-methodology-note

3.	NHS Digital. General Practice Extraction Service (GPES) Data for pandemic planning and research: a guide for analysts and users of the data [Internet]. 2021 [cited 2021 Sep 30];Available from: https://digital.nhs.uk/coronavirus/gpes-data-for-pandemic-planning-and-research/guide-for-analysts-and-users-of-the-data

4.	Ministry of Housing, Communities & Local Government. English indices of deprivation 2019 [Internet]. [cited 2020 Oct 18];Available from: https://www.gov.uk/government/statistics/english-indices-of-deprivation-2019

5.	Digital N. Hospital Episode Statistics (HES) [Internet]. [cited 2021 Oct 6];Available from: https://digital.nhs.uk/data-and-information/data-tools-and-services/data-services/hospital-episode-statistics
