## Supplementary Appendix B for "Trends and associated factors for Covid-19 hospitalisation and fatality risk in 2.3 million adults in England"

Corresponding Author:

Dr Thomas Beaney

**Statistical analysis**

This section provides further detail on the statistical analysis approach. Some of the text here is repeated from the manuscript for clarity.

*Statistical analysis*

Mixed effects logistic regression was used to model the association between each outcome with age and time (model 1) and age and time, along with patient level covariates (model 2), using the *melogit* command in Stata. Clinical Commissioning Group of residence (according to the date of test) was included as a random intercept into all models.

Time was modelled as a restricted cubic spline, representing the week of the Covid-19 test (with week commencing on a Monday) to allow for flexibility in modelling the relationship with the outcomes. A range of number of knots from three to six were considered, with a five-knot spline chosen for analyses based on the Akaike Information Criteria, with knots placed at equally spaced percentiles.^1^

*Multiple imputation*

Multiple imputation using chained equations was used to impute missing values in patient-level covariates sex, ethnicity, IMD decile, BMI category and smoking status. Age was complete in the data set, according to the exclusion criteria. Co-morbidities and Clinically Extremely Vulnerable (CEV) status were assumed to be absent if not recorded, and so by definition included no missing data. Missingness in covariates was assumed to be missing at random, i.e. dependent on the remaining observed data. The *mi impute chained* command in Stata was used, with a logit model for sex and a multinomial logit model for ethnicity, IMD decile, BMI category and smoking status. All variables in the analysis models were also included in the imputation model: age, time splines, age by time spline interactions, CEV status and co-morbidities.^2^

A burn in of 10 iterations was deemed to be sufficient for convergence. Convergence was assessed using a trace plot with 25 iterations, and there was no indication of any trend in the mean or standard deviation of the imputed covariates. In total, 15 imputation sets were created, corresponding roughly to the percentage of missing data in the covariate with most missing values (BMI with 14.0% missing values). In analysis models, the Monte Carlo error of the estimates was consistently less than 10% of the standard errors, indicating satisfactory precision and no requirement for further imputations. Estimates from each imputed data set were combined using Rubin’s rules, applying the Stata *mi estimate* command.

*Predicted probabilities*

For each outcome, the predicted probability of the outcome was calculated within each age group and study week stratum to give age- and time- specific Case Hospitalisation Risk (CHR) and Case Fatality Risk (CFR). These were calculated using the fixed portion of the model (assuming zero random effects). Although using only fixed model components can lead to mis-calibration compared to marginal predictions, simulation studies have shown this to be minimal where the where the intra-class correlation (ICC) is less than 0.15.^3^ The ICC in fully adjusted mixed effects logistic regression models for admission and death were 0.0078 and 0.0087, respectively, suggesting minimal residual variation explained by clustering at CCG level.

Predicted probabilities were calculated for each of the outcome models, and observed probabilities calculated for each decile of predicted probability. Hosmer-Lemeshow plots of deciles of predicted against observed probabilities were plotted. Figures S1 and S2 show the plots for admissions and mortality, respectively in the fully adjusted model (model 2) after imputation. These indicated a linear association between predicted and observed probability and no evidence of mis-calibration of the model.

Analyses were conducted in the Big Data and Analytics Unit Secure Environment, Imperial College. Python v3.9.5 and Pandas v1.2.3 were used in data management. Regression models were conducted in Stata v17.0.

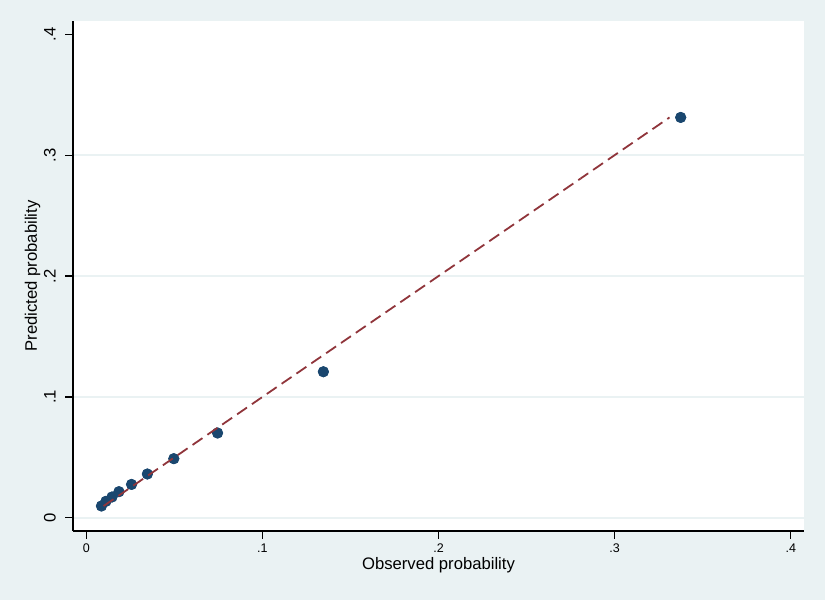
**Figure S1: Hosmer-Lemeshow plot of predicted vs observed probabilities from mixed effects logistic regression model of 28-day emergency admissions, after multiple imputation (N=2,311,282)**

**Figure S2: Hosmer-Lemeshow plot of predicted vs observed probabilities from mixed effects logistic regression model of 28-day mortality, after multiple imputation (N=2,311,282)**

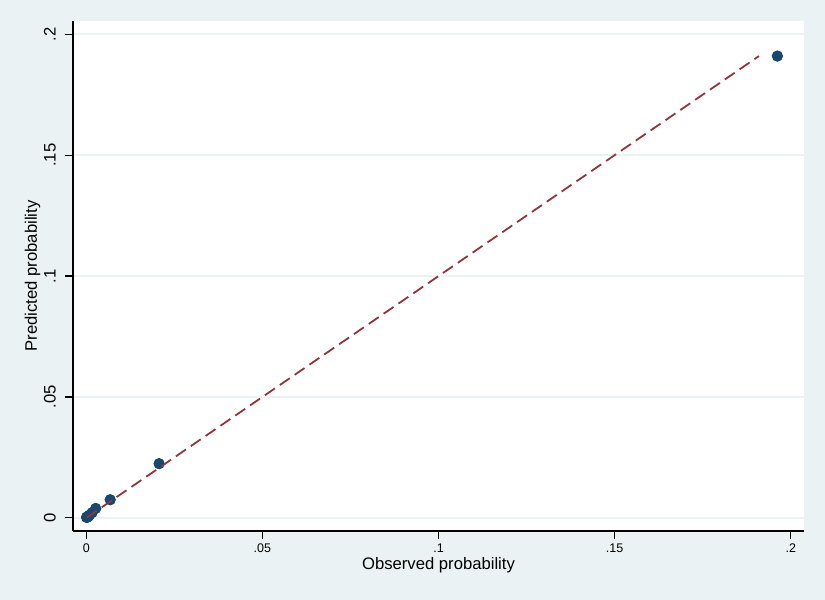

**Table S1: Crude case hospitalisation risk by month of Covid-19 test from October 2020 to April 2021**

| **Age category (years)** | **Hospital admissions within 28 days of positive test** | | | | | | | |
| --- | --- | --- | --- | --- | --- | --- | --- | --- |
|  | **October** | **November** | **December** | **January** | **February** | **March** | **April** | **Total** |
| **18-39** | 1.5% | 1.8% | 1.8% | 2.1% | 2.4% | 2.6% | 2.5% | 2.0% |
| **40-49** | 3.4% | 3.6% | 4.1% | 4.8% | 5.2% | 4.7% | 4.2% | 4.3% |
| **50-59** | 5.4% | 5.7% | 6.9% | 7.8% | 8.2% | 7.4% | 5.8% | 6.9% |
| **60-69** | 10.3% | 11.0% | 12.8% | 14.7% | 16.1% | 13.4% | 8.8% | 13.1% |
| **70-79** | 24.0% | 25.9% | 27.2% | 30.9% | 31.7% | 24.2% | 17.7% | 28.0% |
| **80+** | 38.1% | 37.4% | 38.9% | 41.5% | 39.4% | 38.1% | 39.2% | 39.5% |
| **All ages** | **5.7%** | **6.7%** | **6.8%** | **8.1%** | **8.0%** | **6.1%** | **4.8%** | **7.1%** |

**Table S2: Crude case fatality risk by month of Covid-19 test from October 2020 to April 2021**

| **Age category (years)** | **Deaths within 28 days of positive test** | | | | | | | |
| --- | --- | --- | --- | --- | --- | --- | --- | --- |
|  | **October** | **November** | **December** | **January** | **February** | **March** | **April** | **Total** |
| **18-39** | <0.1% | <0.1% | <0.1% | <0.1% | <0.1% | <0.1% | <0.1% | <0.1% |
| **40-49** | 0.2% | 0.2% | 0.2% | 0.3% | 0.2% | 0.1% | <0.1% | 0.2% |
| **50-59** | 0.4% | 0.6% | 0.7% | 0.9% | 0.9% | 0.5% | 0.4% | 0.7% |
| **60-69** | 2.1% | 2.7% | 3.0% | 3.6% | 3.6% | 2.3% | 1.7% | 3.1% |
| **70-79** | 9.8% | 11.6% | 13.0% | 14.1% | 13.3% | 8.9% | 5.1% | 12.5% |
| **80+** | 28.3% | 30.7% | 34.9% | 34.4% | 30.3% | 22.2% | 20.0% | 32.4% |
| **All ages** | **1.7%** | **2.4%** | **2.3%** | **2.8%** | **2.3%** | **1.1%** | **0.7%** | **2.3%** |

**Table S3: Estimated adjusted odds ratios for emergency hospital admission within 28 days for patient-level predictors from multivariable mixed effects logistic regression models (N=2,311,282)**

| **Outcome** | **Adjusted odds ratio** | **Standard error** | **p-value** | **95% confidence interval** | |
| --- | --- | --- | --- | --- | --- |
|  |  |  |  | **Lower** | **Upper** |
| **Sex** | | | | | |
| Female | Reference | | | | |
| Male | 1.41 | 0.01 | <0.001 | 1.39 | 1.42 |
| **Ethnicity** | | | | | |
| White | Reference | | | | |
| Asian/Asian British | 1.36 | 0.01 | <0.001 | 1.34 | 1.39 |
| Black/African/Caribbean/Black British | 1.40 | 0.02 | <0.001 | 1.36 | 1.44 |
| Mixed/Multiple ethnic groups | 1.32 | 0.03 | <0.001 | 1.26 | 1.39 |
| Other ethnic group | 1.65 | 0.03 | <0.001 | 1.60 | 1.71 |
| **IMD decile** | | | | | |
| 1 (most deprived) | Reference | | | | |
| 2 | 0.93 | 0.01 | 0.54 | 0.91 | 0.95 |
| 3 | 0.87 | 0.01 | 0.001 | 0.85 | 0.89 |
| 4 | 0.84 | 0.01 | 0.001 | 0.82 | 0.86 |
| 5 | 0.82 | 0.01 | <0.001 | 0.80 | 0.84 |
| 6 | 0.77 | 0.01 | <0.001 | 0.75 | 0.79 |
| 7 | 0.74 | 0.01 | <0.001 | 0.72 | 0.76 |
| 8 | 0.73 | 0.01 | <0.001 | 0.71 | 0.75 |
| 9 | 0.71 | 0.01 | <0.001 | 0.69 | 0.73 |
| 10 (least deprived) | 0.68 | 0.01 | <0.001 | 0.66 | 0.70 |
| **Body Mass Index** | | | | | |
| Underweight | 1.10 | 0.02 | <0.001 | 1.05 | 1.14 |
| Healthy weight | Reference | | | | |
| Overweight | 1.24 | 0.01 | <0.001 | 1.22 | 1.26 |
| Obese | 1.93 | 0.02 | <0.001 | 1.90 | 1.97 |
| **Smoking status** | | | | | |
| Never smoker | Reference | | | | |
| Ex-smoker | 1.14 | 0.01 | <0.001 | 1.13 | 1.16 |
| Current smoker | 0.91 | 0.01 | <0.001 | 0.90 | 0.93 |
| **Clinically Extremely Vulnerable** | | | | | |
| No | Reference | | | | |
| Yes | 1.85 | 0.01 | <0.001 | 1.83 | 1.88 |
| **Comorbidities** | | | | | |
| Hypertension | 1.11 | 0.01 | <0.001 | 1.10 | 1.13 |
| Chronic cardiac disease | 1.21 | 0.01 | <0.001 | 1.19 | 1.23 |
| Chronic kidney disease | 1.47 | 0.03 | <0.001 | 1.41 | 1.53 |
| Chronic respiratory disease | 1.30 | 0.01 | <0.001 | 1.28 | 1.32 |
| Dementia | 0.94 | 0.02 | 0.001 | 0.90 | 0.98 |
| Diabetes | 1.33 | 0.01 | <0.001 | 1.31 | 1.35 |
| Chronic neurological disease (including epilepsy) | 1.49 | 0.02 | <0.001 | 1.46 | 1.53 |
| Learning disability | 2.06 | 0.06 | <0.001 | 1.94 | 2.18 |
| Malignancy or immunosuppression | 1.18 | 0.01 | <0.001 | 1.16 | 1.20 |
| Severe mental illness | 1.63 | 0.03 | <0.001 | 1.58 | 1.68 |
| Peripheral vascular disease | 1.17 | 0.02 | <0.001 | 1.13 | 1.22 |
| Stroke or TIA | 1.21 | 0.01 | <0.001 | 1.18 | 1.24 |

Models also included age category and time (study week as a restricted cubic spline with five knots) with an age and time interaction (coefficients not shown). Models adjusted for all other predictors displayed in table.

**Table S4: Estimated adjusted odds ratios for death within 28 days for patient-level predictors from multivariable mixed effects logistic regression models (N=2,311,282)**

| **Outcome** | **Adjusted odds ratio** | **Standard error** | **p-value** | **95% confidence interval** | |
| --- | --- | --- | --- | --- | --- |
|  |  |  |  | **Lower** | **Upper** |
| **Sex** | | | | | |
| Female | Reference | | | | |
| Male | 1.62 | 0.02 | <0.001 | 1.58 | 1.65 |
| **Ethnicity** | | | | | |
| White | Reference | | | | |
| Asian/Asian British | 1.11 | 0.02 | <0.001 | 1.07 | 1.16 |
| Black/African/Caribbean/Black British | 1.14 | 0.04 | <0.001 | 1.07 | 1.22 |
| Mixed/Multiple ethnic groups | 1.02 | 0.06 | 0.712 | 0.91 | 1.15 |
| Other ethnic group | 1.07 | 0.05 | 0.177 | 0.97 | 1.17 |
| **IMD decile** | | | | | |
| 1 (most deprived) | Reference | | | | |
| 2 | 0.94 | 0.02 | 0.005 | 0.90 | 0.98 |
| 3 | 0.89 | 0.02 | <0.001 | 0.85 | 0.93 |
| 4 | 0.86 | 0.02 | <0.001 | 0.83 | 0.90 |
| 5 | 0.80 | 0.02 | <0.001 | 0.77 | 0.84 |
| 6 | 0.79 | 0.02 | <0.001 | 0.75 | 0.82 |
| 7 | 0.75 | 0.02 | <0.001 | 0.72 | 0.79 |
| 8 | 0.72 | 0.02 | <0.001 | 0.69 | 0.75 |
| 9 | 0.71 | 0.02 | <0.001 | 0.68 | 0.75 |
| 10 (least deprived) | 0.70 | 0.02 | <0.001 | 0.67 | 0.74 |
| **Body Mass Index** | | | | | |
| Underweight | 1.99 | 0.06 | <0.001 | 1.87 | 2.11 |
| Healthy weight | Reference | | | | |
| Overweight | 0.80 | 0.01 | <0.001 | 0.77 | 0.82 |
| Obese | 1.04 | 0.02 | 0.012 | 1.01 | 1.07 |
| **Smoking status** | | | | | |
| Never smoker | Reference | | | | |
| Ex-smoker | 1.34 | 0.02 | <0.001 | 1.31 | 1.37 |
| Current smoker | 1.35 | 0.02 | <0.001 | 1.31 | 1.40 |
| **Clinically Extremely Vulnerable** | | | | | |
| No | Reference | | | | |
| Yes | 0.88 | 0.01 | <0.001 | 0.86 | 0.90 |
| **Comorbidities** | | | | | |
| Hypertension | 1.09 | 0.01 | <0.001 | 1.07 | 1.11 |
| Chronic cardiac disease | 1.47 | 0.02 | <0.001 | 1.44 | 1.50 |
| Chronic kidney disease | 2.26 | 0.06 | <0.001 | 2.15 | 2.37 |
| Chronic respiratory disease | 1.38 | 0.02 | <0.001 | 1.35 | 1.41 |
| Dementia | 1.67 | 0.04 | <0.001 | 1.60 | 1.74 |
| Diabetes | 1.44 | 0.02 | <0.001 | 1.41 | 1.47 |
| Chronic neurological disease (including epilepsy) | 1.55 | 0.03 | <0.001 | 1.49 | 1.61 |
| Learning disability | 2.46 | 0.14 | <0.001 | 2.21 | 2.75 |
| Malignancy or immunosuppression | 1.63 | 0.02 | <0.001 | 1.59 | 1.67 |
| Severe mental illness | 2.29 | 0.05 | <0.001 | 2.18 | 2.39 |
| Peripheral vascular disease | 1.42 | 0.03 | <0.001 | 1.35 | 1.48 |
| Stroke or TIA | 1.27 | 0.02 | <0.001 | 1.24 | 1.31 |

Models also include age category and time (study week as a restricted cubic spline with five knots) with an age and time interaction (coefficients not shown). Models adjusted for all other predictors displayed in table.

**Table S5: Estimated adjusted odds ratios for emergency hospital admission within 28 days for patient-level predictors from multivariable mixed effects logistic regression models for the complete cases (N=1,929,999)**

| **Outcome** | **Adjusted odds ratio** | **Standard error** | **p-value** | **95% confidence interval** | |
| --- | --- | --- | --- | --- | --- |
|  |  |  |  | **Lower** | **Upper** |
| **Sex** | | | | | |
| Female | Reference | | | | |
| Male | 1.44 | 0.01 | <0.001 | 1.42 | 1.46 |
| **Ethnicity** | | | | | |
| White | Reference | | | | |
| Asian/Asian British | 1.37 | 0.01 | <0.001 | 1.34 | 1.39 |
| Black/African/Caribbean/Black British | 1.40 | 0.02 | <0.001 | 1.36 | 1.44 |
| Mixed/Multiple ethnic groups | 1.32 | 0.03 | <0.001 | 1.26 | 1.38 |
| Other ethnic group | 1.67 | 0.03 | <0.001 | 1.61 | 1.73 |
| **IMD decile** | | | | | |
| 1 (most deprived) | Reference | | | | |
| 2 | 0.94 | 0.01 | <0.001 | 0.92 | 0.96 |
| 3 | 0.88 | 0.01 | <0.001 | 0.86 | 0.90 |
| 4 | 0.84 | 0.01 | <0.001 | 0.82 | 0.86 |
| 5 | 0.83 | 0.01 | <0.001 | 0.81 | 0.85 |
| 6 | 0.78 | 0.01 | <0.001 | 0.76 | 0.80 |
| 7 | 0.75 | 0.01 | <0.001 | 0.73 | 0.77 |
| 8 | 0.75 | 0.01 | <0.001 | 0.73 | 0.77 |
| 9 | 0.72 | 0.01 | <0.001 | 0.70 | 0.74 |
| 10 (least deprived) | 0.70 | 0.01 | <0.001 | 0.68 | 0.72 |
| **Body Mass Index** | | | | | |
| Underweight | 1.12 | 0.02 | <0.001 | 1.08 | 1.17 |
| Healthy weight | Reference | | | | |
| Overweight | 1.25 | 0.01 | <0.001 | 1.23 | 1.27 |
| Obese | 1.95 | 0.02 | <0.001 | 1.92 | 1.98 |
| **Smoking status** | | | | | |
| Never smoker | Reference | | | | |
| Ex-smoker | 1.14 | 0.01 | <0.001 | 1.12 | 1.16 |
| Current smoker | 0.91 | 0.01 | <0.001 | 0.90 | 0.93 |
| **Clinically Extremely Vulnerable** | | | | | |
| No | Reference | | | | |
| Yes | 1.86 | 0.02 | <0.001 | 1.83 | 1.89 |
| **Comorbidities** | | | | | |
| Hypertension | 1.17 | 0.01 | <0.001 | 1.15 | 1.18 |
| Chronic cardiac disease | 1.24 | 0.01 | <0.001 | 1.22 | 1.26 |
| Chronic kidney disease | 1.46 | 0.03 | <0.001 | 1.40 | 1.51 |
| Chronic respiratory disease | 1.34 | 0.01 | <0.001 | 1.32 | 1.35 |
| Dementia | 0.98 | 0.02 | 0.294 | 0.94 | 1.02 |
| Diabetes | 1.34 | 0.01 | <0.001 | 1.32 | 1.36 |
| Chronic neurological disease (including epilepsy) | 1.50 | 0.02 | <0.001 | 1.46 | 1.54 |
| Learning disability | 2.05 | 0.06 | <0.001 | 1.94 | 2.18 |
| Malignancy or immunosuppression | 1.20 | 0.01 | <0.001 | 1.18 | 1.22 |
| Severe mental illness | 1.64 | 0.03 | <0.001 | 1.59 | 1.69 |
| Peripheral vascular disease | 1.18 | 0.02 | <0.001 | 1.14 | 1.23 |
| Stroke or TIA | 1.23 | 0.02 | <0.001 | 1.20 | 1.26 |

Models also included age category and time (study week as a restricted cubic spline with five knots) with an age and time interaction (coefficients not shown). Models adjusted for all other predictors displayed in table.

**Table S6: Estimated adjusted odds ratios for death within 28 days for patient-level predictors from multivariable mixed effects logistic regression models for the complete cases ((N=1,929,999))**

| **Outcome** | **Adjusted odds ratio** | **Standard error** | **p-value** | **95% confidence interval** | |
| --- | --- | --- | --- | --- | --- |
|  |  |  |  | **Lower** | **Upper** |
| **Sex** | | | | | |
| Female | Reference | | | | |
| Male | 1.62 | 0.02 | <0.001 | 1.58 | 1.65 |
| **Ethnicity** | | | | | |
| White | Reference | | | | |
| Asian/Asian British | 1.10 | 0.02 | <0.001 | 1.06 | 1.15 |
| Black/African/Caribbean/Black British | 1.13 | 0.04 | <0.001 | 1.06 | 1.21 |
| Mixed/Multiple ethnic groups | 1.02 | 0.06 | 0.747 | 0.91 | 1.15 |
| Other ethnic group | 1.06 | 0.05 | 0.235 | 0.96 | 1.16 |
| **IMD decile** | | | | | |
| 1 (most deprived) | Reference | | | | |
| 2 | 0.95 | 0.02 | 0.021 | 0.91 | 0.99 |
| 3 | 0.90 | 0.02 | <0.001 | 0.86 | 0.94 |
| 4 | 0.88 | 0.02 | <0.001 | 0.84 | 0.92 |
| 5 | 0.81 | 0.02 | <0.001 | 0.77 | 0.85 |
| 6 | 0.81 | 0.02 | <0.001 | 0.77 | 0.84 |
| 7 | 0.76 | 0.02 | <0.001 | 0.73 | 0.80 |
| 8 | 0.75 | 0.02 | <0.001 | 0.71 | 0.78 |
| 9 | 0.72 | 0.02 | <0.001 | 0.69 | 0.76 |
| 10 (least deprived) | 0.72 | 0.02 | <0.001 | 0.69 | 0.76 |
| **Body Mass Index** | | | | | |
| Underweight | 1.98 | 0.06 | <0.001 | 1.86 | 2.10 |
| Healthy weight | Reference | | | | |
| Overweight | 0.81 | 0.01 | <0.001 | 0.78 | 0.83 |
| Obese | 1.05 | 0.01 | 0.001 | 1.02 | 1.08 |
| **Smoking status** | | | | | |
| Never smoker | Reference | | | | |
| Ex-smoker | 1.34 | 0.02 | <0.001 | 1.31 | 1.37 |
| Current smoker | 1.35 | 0.02 | <0.001 | 1.31 | 1.40 |
| **Clinically Extremely Vulnerable** | | | | | |
| No | Reference | | | | |
| Yes | 0.89 | 0.01 | <0.001 | 0.87 | 0.92 |
| **Comorbidities** | | | | | |
| Hypertension | 1.18 | 0.01 | <0.001 | 1.16 | 1.21 |
| Chronic cardiac disease | 1.55 | 0.02 | <0.001 | 1.52 | 1.59 |
| Chronic kidney disease | 2.25 | 0.06 | <0.001 | 2.14 | 2.36 |
| Chronic respiratory disease | 1.45 | 0.02 | <0.001 | 1.42 | 1.49 |
| Dementia | 1.76 | 0.04 | <0.001 | 1.68 | 1.84 |
| Diabetes | 1.49 | 0.02 | <0.001 | 1.45 | 1.52 |
| Chronic neurological disease (including epilepsy) | 1.59 | 0.03 | <0.001 | 1.53 | 1.66 |
| Learning disability | 2.48 | 0.14 | <0.001 | 2.22 | 2.77 |
| Malignancy or immunosuppression | 1.71 | 0.02 | <0.001 | 1.67 | 1.75 |
| Severe mental illness | 2.35 | 0.06 | <0.001 | 2.25 | 2.46 |
| Peripheral vascular disease | 1.42 | 0.03 | <0.001 | 1.36 | 1.49 |
| Stroke or TIA | 1.30 | 0.02 | <0.001 | 1.26 | 1.34 |

Models also include age category and time (study week as a restricted cubic spline with five knots) with an age and time interaction (coefficients not shown). Models adjusted for all other predictors displayed in table.

**Figure S3: Case hospitalisation risk (A) and fatality risk (B) over time in people with Covid-19 from unadjusted mixed effects logistic regression models (N=2,311,282)**

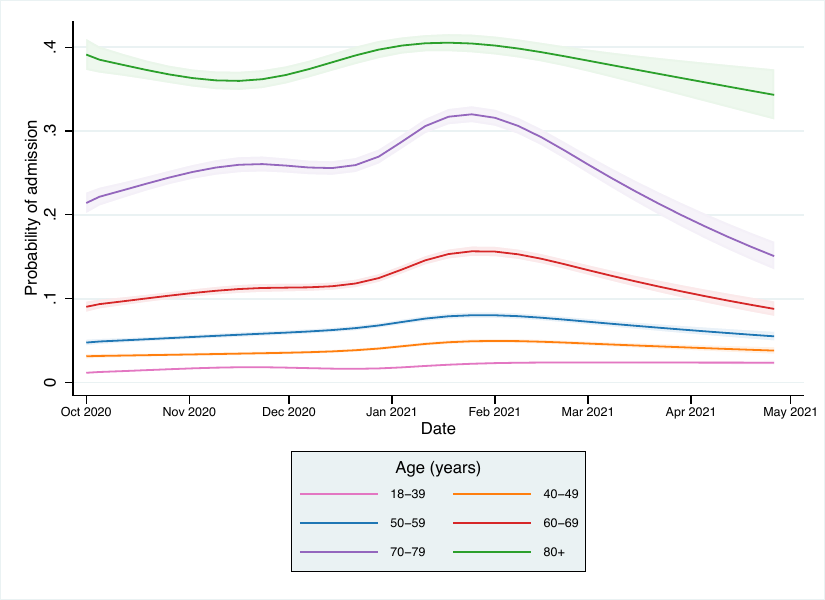

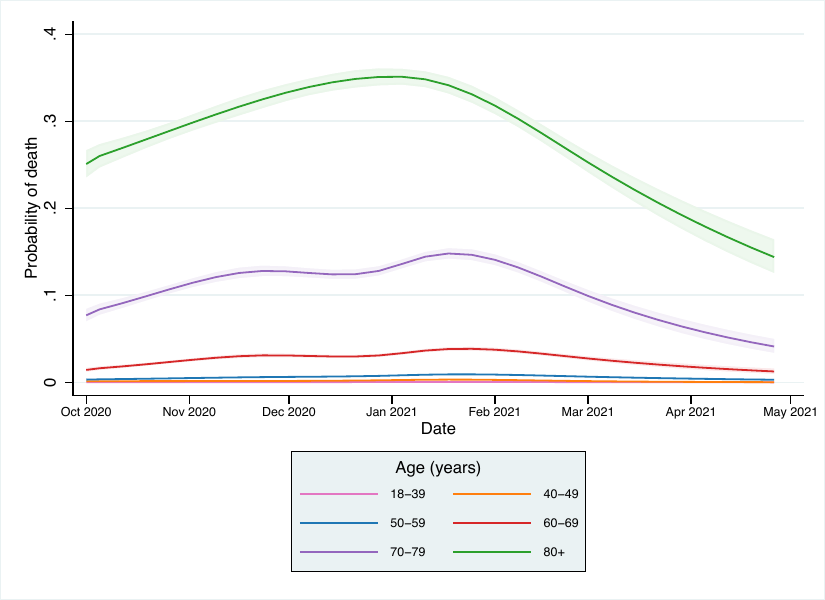

**A**

**B**

**Figure S4: Relative probability of hospital admission within 28 days over time in people with Covid-19 from unadjusted mixed effects logistic regression models (N=2,311,282)**

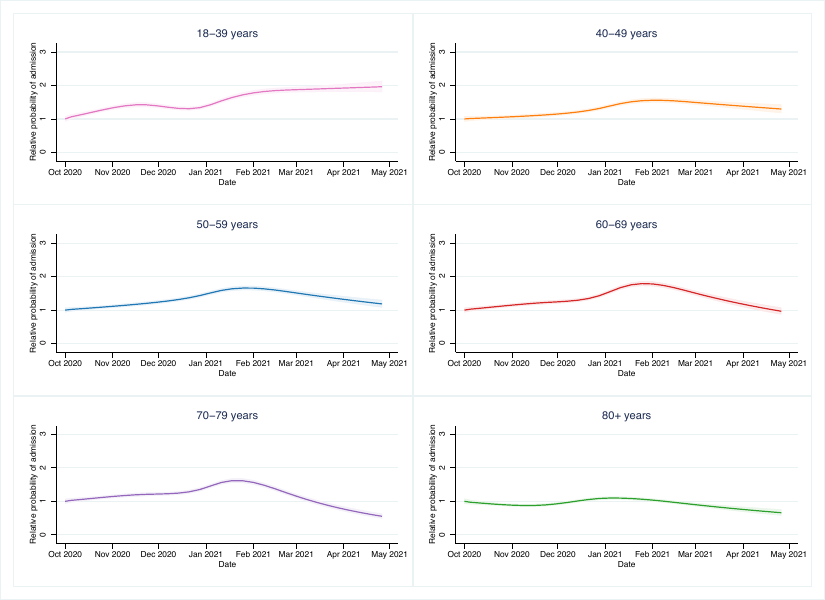

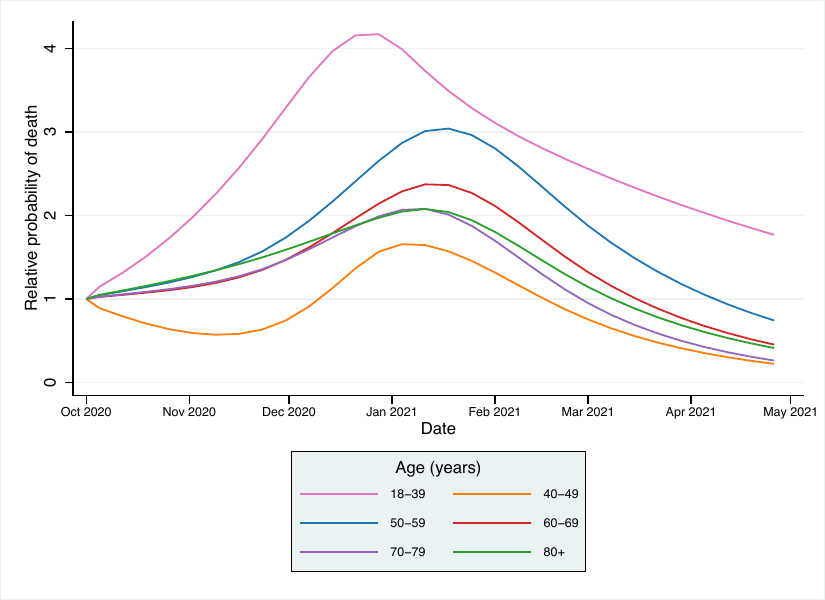

**Figure S5: Relative probability of death with 28 days over time in people with Covid-19 from unadjusted mixed effects logistic regression models (N=2,311,282)**

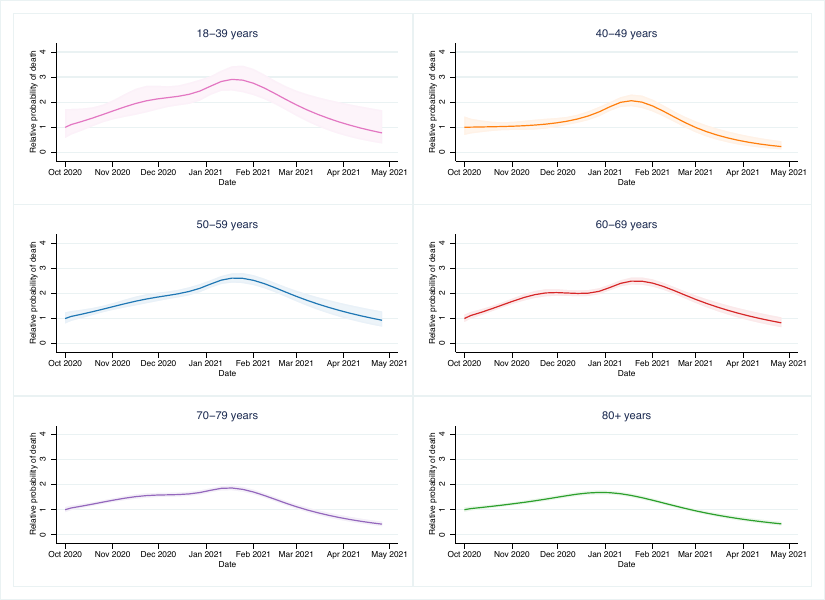

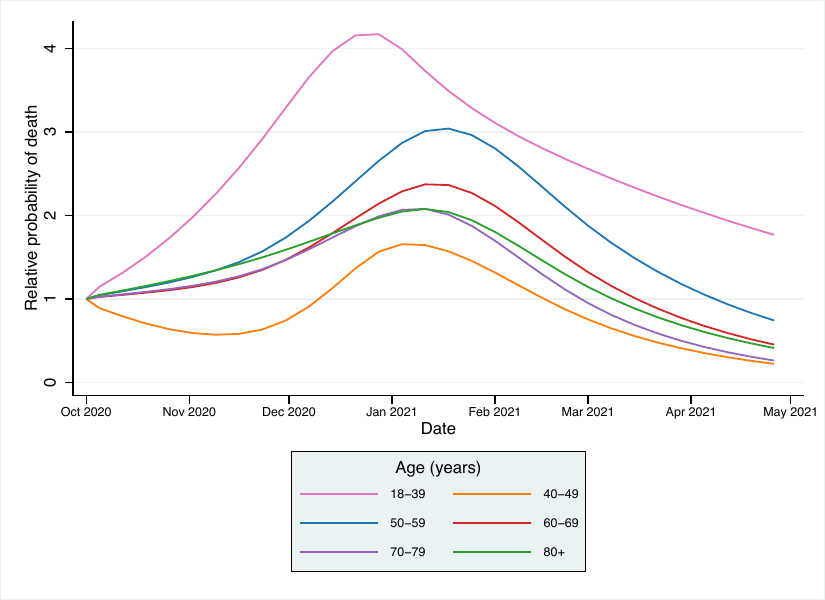
